# The Approved Dose of Ivermectin Alone is not the Ideal Dose for the Treatment of COVID-19

**DOI:** 10.1101/2020.04.21.20073262

**Authors:** Virginia D. Schmith, Jie (Jessie) Zhou, Lauren RL Lohmer

## Abstract

**Introduction:** Caly, Druce (1) reported that ivermectin inhibited SARS-CoV-2 *in vitr*o for up to 48 h using ivermectin at 5μM. The concentration resulting in 50% inhibition (IC_50,_ 2 µM) was >35x higher than the maximum plasma concentration (Cmax) after oral administration of the approved dose of ivermectin when given fasted.

**Method:** Simulations were conducted using an available population pharmacokinetic model to predict total (bound and unbound) and unbound plasma concentration-time profiles after a single and repeat fasted administration of the approved dose of ivermectin (200 μg/kg), 60 mg, and 120 mg. Plasma total Cmax was determined and then multiplied by the lung:plasma ratio reported in cattle to predict the lung Cmax after administration of each single dose.

**Results:** Plasma ivermectin concentrations of total (bound and unbound) and unbound concentrations do not reach the IC_50_, even for a dose level 10x higher than the approved dose. Even with higher exposure in lungs than plasma, ivermectin is unlikely to reach the IC_50_ in lungs after single oral administration of the approved dose (predicted lung: 0.0857 µM) or at doses 10x higher that the approved dose administered orally (predicted lung: 0.817 µM).

**Conclusions:** The likelihood of a successful clinical trial using the approved dose of ivermectin is low. Combination therapy should be evaluated *in vitro*. Re-purposing drugs for use in COVID-19 treatment is an ideal strategy but is only feasible when product safety has been established and experiments of re-purposed drugs are conducted at clinically relevant concentrations.

## Introduction

Recently, an article by Caly, Druce (1) reported that ivermectin inhibited SARS-CoV-2 *in vitr*o causing a ∼5000-fold reduction in viral RNA at 48 h with ivermectin at 5 μM. The concentration resulting in 50% inhibition (IC_50_) of 2 μM (1750 ng/mL) is >35x higher than the maximum plasma concentration (Cmax) of 0.05 µM (46.6 ng/mL (2)) after oral administration of the approved dose (∼200 μg/kg). Since ivermectin has high protein binding (93%, (3)), the IC_50_ is orders of magnitude higher than the unbound plasma Cmax after approved doses of ivermectin (0.0035 µM; 3.26 ng/mL).

In order to understand how *in vitro* SARS-CoV-2 inhibition by ivermectin translates to humans, one must first evaluate these concentrations compared to predicted lung concentrations in humans after oral administration of ivermectin. Ivermectin reaching the lung after oral dosing is likely related to the unbound concentrations in plasma, the lipophilicity, and any transporters that may help maintain tissue distribution. While ivermectin concentrations in lung tissue cannot be measured in humans, ivermectin lung exposure was reported to be 2.7x higher than total plasma exposure in cattle after a single dose (4). Even with higher concentrations in lung, ivermectin is unlikely to reach the IC_50_ after oral administration of the approved dose in humans.

Unlike the narrow therapeutic index for hydroxychloroquine and chloroquine, ivermectin has a wider safety margin (2). The safety of higher doses of ivermectin has been evaluated in a Phase 3 study, where 200-400 μg/kg doses were studied in patients with Dengue fever (5, 6). Even higher doses (up to 10x higher than approved doses) were studied in a small Phase 1 trial (7). This trial showed that ivermectin administered orally in the fasted state was well tolerated both after a single 120 mg dose (10x higher than approved dose) and after 60 mg three times weekly (every 72 hours). Adverse experiences were similar between ivermectin and placebo and did not increase with dose. All dosing regimens had a mydriatic effect (the primary safety endpoint based on results from toxicology studies) similar to placebo. It is important to note that while this study evaluated common adverse events, the presence or incidence of rare adverse events at these high doses are unknown, given the small number of subjects studied.

The overall objective of this analysis was to evaluate what doses in humans would be predicted to result in lung concentrations reaching the IC_50_ in the lungs to help in designing a successful clinical trial with ivermectin in the treatment of COVID-19.

## Methods

A population pharmacokinetic model for ivermectin reported by Duthaler, Suenderhauf (8) was used in the simulations. This model was a two-compartment model with a transit absorption model, first-order elimination, and weight as a covariate on central volume of distribution and clearance. This model was developed from healthy subjects receiving a single ivermectin dose of 12 mg in the fed state.

Simulations (n=100) were performed using NONMEM version 7.4 (ICON). Total (bound and unbound) plasma concentration-time profiles were simulated to predict exposure for the approved dose of ivermectin (200 μg/kg, in 3 mg increments) and 120 mg (studied by Guzzo, Furtek (7) as single doses). Since ivermectin concentrations remained steady in cattle lungs for 8 days and then declined over an additional 30 days after a single subcutaneous dose (4), additional simulations were conducted to predict plasma concentrations with weekly dosing. In addition, 60 mg administered three times weekly was simulated (every 72 hours), given this dose was studied in healthy subjects by Guzzo, Furtek (7).

Body weights were sampled from Center for Disease Control weight chart assuming 20 year old adults, with male:female ratio being 1:1 (9).

Because the population pharmacokinetic model was built based on subjects who received ivermectin with a high-fat breakfast, bioavailability is increased by 2.57-fold increase in fed state with no change in Tmax (7), and ivermectin should be taken on an empty stomach with water (2), all plasma concentration-time data were divided by the geometric least squares mean ratio fed:fasted (2.57) to predict concentrations when ivermectin is administered in the fasted state.

Unbound plasma concentration-time data were predicted by multiplying the total concentration by the unbound fraction (0.068).

The Cmax values for total plasma concentration were determined and multiplied by the lung:plasma ratio (2.67:1) in cattle reported by Lifschitz, Virkel (4) at each single dose to derive the Cmax values for total lung concentrations. The lung:plasma ratio after repeat dosing could not be determined without further modeling of the data from cattle. Some accumulation is expected in lung (but not plasma) with weekly or 3x weekly administration, but needs further investigation with more experimental data. A ball-park accumulation ratio (AR) in lung was calculated using Equation 1: 

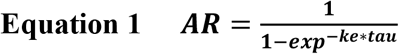

Where ke is the estimated elimination rate constant from the cattle lung concentration-time profile presented by Lifschitz, Virkel (4) and tau is the dosing interval.

## Results

Plasma ivermectin concentrations of total (bound and unbound) and unbound concentrations do not reach the IC_50_ reported by Caly, Druce (1), even for dose level 10x higher than the approved dose, or after repeat dosing (Figure 1). Plasma exposures did not increase substantially after repeat dosing, with very limited ivermectin accumulation in plasma after 3 times weekly or weekly dosing.

**Figure 1.**
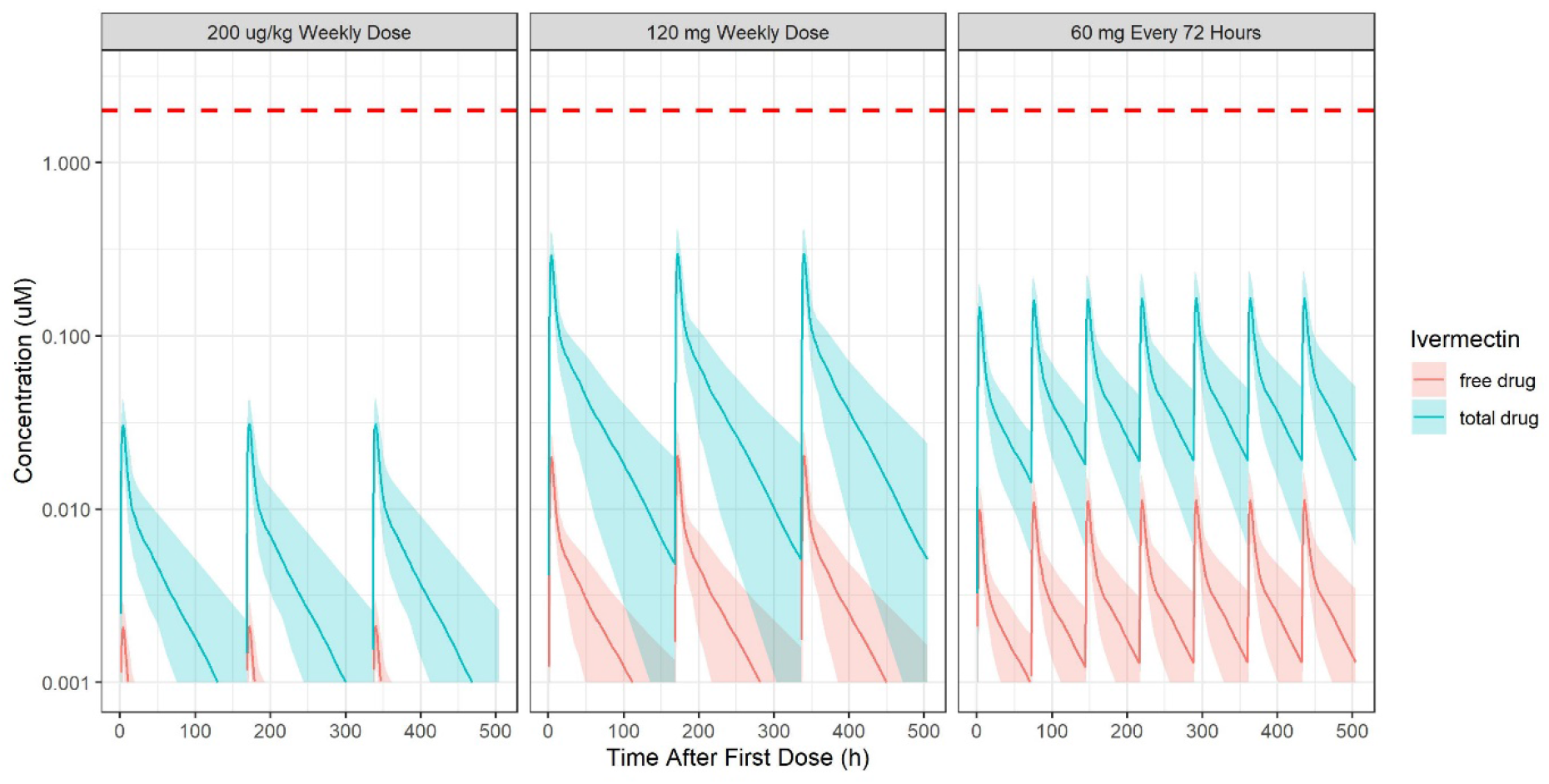
Total (Bound and Unbound) and Unbound Plasma Ivermectin Concentrations Over Time Relative to the IC_50_ After Approved Doses of Ivermectin (200 µg/kg) Administered Once Weekly (instead of as a Single Dose), and After Higher Doses Including 60 mg Every Three Days or 120 mg Once Weekly Red dashed line=IC_50_ reported by Caly et al. (2020), 2 µM (1750 ng/mL). Blue shaded area and line=total plasma drug concentration and its 95% confidence interval (CI); red shaded area and line=free plasma drug concentration and its 95% CI.

Even with higher exposure in the lung, ivermectin is unlikely to reach the IC_50_ of 2 µM in the lung after single oral administration of the approved dose (predicted lung concentration: 0.0857 µM) or at doses 10x higher that the approved dose administered orally (predicted lung concentration: 0.817 µM) (Table 1). Currently there is no lung tissue disposition data available after repeated dosing, but the IC_50_ would not be reached in lungs unless there was >25-fold accumulation in lungs with ivermectin at the approved dose administered weekly, >2.5-fold for 120 mg administered once weekly, or >5-fold after ivermectin 60 mg every 72 hours. If the approved dose was administered three times weekly, the ball-park accumulation ratio in lung tissue is 2.20, which would result in lung concentrations that are 1/10^th^ of the IC_50_. If the approved dose was administered daily, the ball-park accumulation ratio in lung tissue is 5.35, which would result in lung concentrations that are 1/4^th^ of the IC_50_.

**Table 1.** Predicted Maximum Total Plasma Concentrations and Lung Concentrations After Various Doses of Ivermectin Administered Fasted.

| Treatment |  | Predicted Total Cmax (µM)<br>Median [2.5<sup>th</sup>, 97.5<sup>th</sup> Percentiles] |  |
| --- | --- | --- | --- |
|  |  | Plasma | Lung<sup>b</sup> |
| Single Dose | 200 µg/kg Single Dose<br>(Labelled Dose) | 0.0321<br>[0.0234, 0.0444] | 0.0857<br>[0.0625, 0.119] |
|  | 120 mg Single Dose<sup>a</sup> | 0.306<br>[0.213, 0.411] | 0.817<br>[0.569, 1.10] |
| Repeated Dose<br>(3 Weeks) | 200 µg/kg Weekly | 0.0306<br>[0.0213, 0.0411] |  |
|  | 60 mg Every 72 hours<sup>a</sup> | 0.173<br>[0.108, 0.240] |  |
|  | 120 mg Weekly | 0.312<br>[0.215, 0.430] |  |

<sup>a</sup>Each Administered to 12 subjects (Guzzo et al., 2002)
<sup>b</sup>Calculated based on reported lung: plasma ratio of 2.67 in cattle (Lifschitz et al., 2000).
CI = Confidence Interval

## Discussion

The *in vitro* studies showing that ivermectin inhibited SARS-CoV-2 (1) were conducted at concentrations that were substantially higher than predicted plasma and lung concentrations in humans receiving the approved dose of ivermectin. Therefore, the likelihood of a successful clinical trial using the approved dose of ivermectin is low. If a clinical trial is conducted, a well-controlled dose-response study should be considered and the feasibility of ivermectin as an inhaled treatment should be evaluated.

A first step would be to repeat the *in vitro* study reported by Caly, Druce (1) with other compounds that could theoretically potentiate ivermectin’s inhibition of SARS-COV-2 to identify whether a concentration of 0.1 μM (rather than 5 μM) could inhibit SARS-COV-2. Re-purposing drugs for use in the treatment of COVID-19 is an ideal strategy but is only feasible if the safety of the product use has been established at the dose levels that produce efficacy. Therefore, *in vitro* experiments of re-purposed drugs should be conducted at clinically relevant concentrations.

As soon as the *in vitro* findings were published, clinicians all over the world were using ivermectin off-label. Patel et al. reported this week through an observational registry-based study from 169 hospitals across the world that ivermectin 150 μg/kg administered to 52 patients after institution of mechanical ventilation showed a potential decrease in hospital stay length and survival benefit compared to 1918 conventionally-treated patients (10). It is noted that these results did not account for co-morbidities that might account for these differences. Nevertheless, if ivermectin did contribute to these clinical findings, it would suggest that the *in vitro* findings by Caly, Druce (1) do not correlate with the very small amounts of the drug at the site of action in humans, concentrations of drug do not need to reach the IC_50_ for clinical benefit, distribution into the lungs of humans is greater than in cattle, or that accumulation in lung tissue is much greater (>25-fold) than expected after repeat dosing.

If a clinical study is conducted with ivermectin, it would be important to conduct a well-controlled clinical dose-response study with ivermectin at a low dose (the approved dose, with lower likelihood of success) and at a higher dose relative to placebo in patients with COVID-19. The ideal higher dose of ivermectin has not been established. Ivermectin doses up to 120 mg have only been administered to a small number of subjects (7). After daily dosing of the ivermectin 200 μg/kg, lung concentrations are predicted to be around 1/4^th^ of the IC_50_. Daily dosing of ivermectin at the approved dose for longer periods (e.g., 14 days) have only been studied in small studies for serious infections where an unapproved subcutaneous formulation was used (11). Therefore, if higher doses are studied weekly, if the approved dose is studied daily, or a parenteral formulation is used, subjects will need to be monitored closely. In addition, the study would need to control for factors affecting variability in the exposure to ivermectin including administering the dose in the fasted state and excluding P-glycoprotein and CYP3A4 inhibitors (2).

Lastly, a potential longer-term solution would be to consider whether inhaled treatment with ivermectin is feasible. Inhaled treatment would allow for higher concentrations at the site of action while limiting the systemic exposure but may require further study of the safety and tolerability in animals prior to human exposure. Only one nonclinical study has been published on inhaled ivermectin, in which the no-observed-adverse-effect level (NOAEL) after 28 days of inhaled ivermectin was identified to be 380 mg/m^3^ (12), and no studies using the inhaled route of administration have been identified in humans. Of key importance is determining whether ivermectin has general properties that would allow inhalation, with no local tolerability issues. Experts must, therefore, evaluate whether ivermectin possesses the ideal properties for inhalation, and whether inhalation of ivermectin poses any theoretical risks that might limit this route of administration.

Overall, the results identified in the paper by Caly, Druce (1) create an opportunity for interdisciplinary collaboration in helping to understand the highest probability of success for ivermectin treatment, prior to exploration in clinical studies (or worse yet, off-label use by the general public) with a less-than-ideal dose.

## Data Availability

This research does not involve the administration of treatment to patients, but does involve simulation of patient data. The simulations are available upon request.

## Study Highlights

### What is the current knowledge on the topic?

Caly, Druce (1) reported that ivermectin inhibited SARS-CoV-2 *in vitr*o with an IC_50_ which was >35x higher than the Cmax after oral administration of the approved dose of ivermectin.

### What question did this study address?

What ivermectin dose reaches the IC_50_ in lungs after oral administration in humans.

### What does this study add to our knowledge?

Ivermectin is unlikely to reach the IC_50_ in lungs after oral administration of the approved dose or doses 10x higher than the approved doses as a single dose. The approved dose of ivermectin alone has a low probability of a success in the treatment of COVID-19.

### How might this change clinical pharmacology or translational science?

Re-purposing drugs for use in COVID-19 treatment is an ideal strategy but is only feasible when product safety has been established and experiments of re-purposed drugs are conducted at clinically relevant concentrations.

## Acknowledgements

The authors would like to thank Anginelle Alabanza, MS, RAC, for her help reviewing and editing the manuscript.

## Author Contributions

GS wrote the manuscript and designed the research. JZ wrote the manuscript, performed the research, and analyzed the data. LL wrote the manuscript and analyzed the data.

## Notes

### Competing Interest Statement

VS, LL, and JZ are consultants and have worked with many companies but have not worked with companies who produce ivermectin orally (EDENBRIDGE PHARMS and MERCK). VS has previously received a grant from the Bill and Melinda Gates foundation unrelated to this work.

### Funding Statement

This research was not funded

## References

(1) Caly, L., Druce, J., Catton, M., Jans, D. & Wagstaff, K. The FDA-approved Drug Ivermectin inhibits the replication of SARS-CoV-2 in vitro. Antivir Res Pre-proof, (2020).

(2) STROMECTOL® (ivermectin) tablets [package insert]. Merck Sharp & Dohme Corp. <https://www.merck.com/product/usa/pi_circulars/s/stromectol/stromectol_pi.pdf> (Rev. 2018).

(3) Klotz, U., Ogbuokiri, J.E. & Okonkwo, P.O. Ivermectin binds avidly to plasma proteins. European journal of clinical pharmacology 39, 607–8 (1990).

(4) Lifschitz, A. et al. Comparative distribution of ivermectin and doramectin to parasite location tissues in cattle. Veterinary parasitology 87, 327–38 (2000).

(5) NCT02045069. Efficacy and Safety of Ivermectin Against Dengue Infection. (https://clinicaltrials.gov/ct2/show/NCT02045069, 2015).

(6) (2018). Efficacy and Safety of Ivermectin against Dengue Infection: A Phase III, Randomized, Double-blind, Placebo-controlled Trial. The 34th Annual Meeting The Royal College of Physicians of Thailand.

(7) Guzzo, C.A. et al. Safety, tolerability, and pharmacokinetics of escalating high doses of ivermectin in healthy adult subjects. Journal of clinical pharmacology 42, 1122–33 (2002).

(8) Duthaler, U. et al. Population pharmacokinetics of oral ivermectin in venous plasma and dried blood spots in healthy volunteers. British journal of clinical pharmacology 85, 626–33 (2019).

(9) Centers for Disease Control and Prevention. (2009). United States Growth Charts [dataset] (National Center for Health Statistics, 2009).

(10) Patel, A. & Desai, S. Ivermectin in COVID-19 Related Critical Illness (April 6, 2020). Available at SSRN: https://ssrncom/abstract=3570270 or http://dxdoiorg/102139/ssrn3570270, (2020).

(11) Turner, S.A., Maclean, J.D., Fleckenstein, L. & Greenaway, C. Parenteral administration of ivermectin in a patient with disseminated strongyloidiasis. The American journal of tropical medicine and hygiene 73, 911–4 (2005).

(12) Ji, L. et al. Study on the subacute inhalation toxicity of ivermectin TC in rats. Chinese Journal of Comparative Medicine 26, 70–4 (2016).

